# Data to Practice (D2P). Protocol for the development, dissemination and initial implementation of best practice guides for common musculoskeletal conditions: a mixed-methods study

**DOI:** 10.64898/2026.04.09.26350486

**Authors:** Dylan Morrissey, Faiza Sharif, Angie Fearon, Bradley Stephen Neal, Tobias Bremer, Paul Swinton, Phillip Newman, Simon Lack, Kay Cooper, Rodrigo Rabello, D2P Group

## Abstract

**Introduction:** Musculoskeletal conditions have high, and increasing, incidence and prevalence. Although there are many clinical guidelines available for common conditions, most are poor quality and sparsely adopted into practice. We aim to improve patient outcomes by developing robust Best Practice Guidelines (BPG) to get research findings into practice for a range of common musculoskeletal conditions.

**Methods and analysis:** Mixed methods with systematic review of high-quality studies and qualitative elicitation of both patient’s perspectives and expert clinical reasoning through in-depth interviews will form the basis for the BPGs. A segregated convergent synthesis, informed throughout by stakeholder engagement, will guide the format and structure of the BPGs.

**Ethics, outputs and dissemination:** Ethical approval for the qualitative studies and implementation events will be obtained from university and health service research ethics committees. Educational packages for each BPG condition will be hosted online and be available for students, clinicians, and education providers. Dissemination will follow traditional routes including publications and presentations; alongside innovative approaches such as collaboration with higher education institutions, online hosting, adoption by professional bodies, and a social media campaign. Implementation will occur adaptively in multiple national contexts to reflect local requirements and resources, deploying participatory and implementation methods that are contextually and culturally appropriate.

**KEY MESSAGES:**

- **What is already known on this topic** – Clinical guidelines for the management of musculoskeletal conditions are common, but have limitations regarding quality, applicability, editorial independence, and patient perspective. They are rarely adopted into clinical practice.
- **What this study adds** – We have developed a robust (supported by Patient and Participant Involvement) mixed-methods approach that integrates the three components of evidence-based medicine: synthesis of high-quality evidence, patients’ perspectives/values, and expert clinical reasoning. We have also developed an education, dissemination, and implementation approach to facilitate international adoption of these guidelines.
- **How this study might affect research, practice or policy** – The guideline development methods will integrate the three pillars of evidence-based practice and ensure they are robust and clinically applicable. Creation of educational material combined with an implementation and dissemination plan will support adoption into clinical practice of different countries and cultures, designed to lead to improved patient outcomes.

## BACKGROUND

Musculoskeletal (MSK) conditions are the most common contributor to disability across the world. Approximately 1.71 billion people of all ages live with musculoskeletal conditions, accounting for 17% of all years lived with disability.^1^ The 2023 Versus Arthritis report on the state of musculoskeletal health in the United Kingdom (UK) details that persistent musculoskeletal conditions are common; with one third of the UK population (>20 million people) currently living with at least one.^2^ They have high societal impact, being the third largest area of NHS spending, costing an estimated £6.3 billion between 2022-2023.^2^

Musculoskeletal conditions continue to impose a substantial burden on individuals, with many experiencing long-term symptoms despite available care. For example, 65% of people with low back pain are still symptomatic after 12 months,^3^ 45% of people with greater trochanteric pain syndrome still present with symptoms after 11 years,^4^ over half of people with patellofemoral pain are still symptomatic five years post-treatment,^5^ and 46% of people with plantar heel pain are symptomatic after 5 to 27 years.^6^

These are all conditions for which there should be better outcomes. For example, at 12 months, people with gluteal tendinopathy showed 78% improvement with education and exercise,^7^ those with patellofemoral pain showed 73–84% improvement with foot orthoses or physiotherapy,^8^ those with low-back pain improved from 9.8 to 5.4 on the Roland and Morris Disability Questionnaire (24-point scale) with the STarT Back protocol,^9^ and those with plantar heel pain achieved at least 60% pain relief in 88% of cases with radial shockwave therapy.^10^

There are numerous reasons why this mismatch occurs, with the difficulty in assimilating the explosion of evidence into practice being key. We argue that the primary fault is in the lack of high-quality, accessible, meaningful evidence syntheses that are strongly evidence-based, incorporate clear clinical reasoning, and prioritise the patient perspective to be contextually relevant. A 2019 publication also reports that where they exist, close to 50% of clinicians failed to adhere to evidence-informed guidelines but without providing information as to why that may be.^11^ Our presumption is that guidelines would be better followed, and outcomes improved accordingly, with better dissemination, which has previously been proven this for common conditions.^12^ Projections are that the prevalence and societal impact of musculoskeletal conditions will continue to rise,^1^ making it imperative that high-quality, evidence-informed care be defined and consistently provided.

Methods for developing best practice guides (BPGs) exist to improve the standard of healthcare provision.^13^ Although several guidelines have been developed to guide the management of several conditions, the majority are poor quality and typically lack applicability, editorial independence, and the patient perspective.^14^ We have developed a mixed methods approach to producing BPGs by synthesising systematic review with meta-analysis, the lived experience of patients, and the clinical reasoning of experts for people with plantar heel pain^15^ and for people with patellofemoral pain.^16^ This protocol is for development of BPGs for a further six common musculoskeletal conditions, in parallel with adaptive implementation and dissemination packages for all 8.

Our aim is to guide care, disseminate effectively, and ultimately improve patient outcomes. We will apply and extend our established mixed-methods approach to producing these BPGs, integrating evidence of effectiveness, patients’ lived experience, and expert clinical reasoning while following the Appraisal of Guidelines, Research and Evaluation tool (AGREE II). Throughout the project, we will engage key stakeholders, conduct rigorous systematic reviews of high-quality trials, identify key aspects of patient perspectives on care, elicit the clinical reasoning of inter-disciplinary world experts using qualitative methods, and synthesise the findings into multiple formats to support dissemination and early implementation. The impact of success would be improved evidence translation, reduced variability of care, and better long-term patient-reported outcomes.

## METHODS

### BPG Design

This protocol is reported adhering to the Preferred Reporting Items for Systematic Review and Meta-Analysis Protocols (PRISMA-P).^17^ The RAiSE (Responsible use of AI in evidence SynthEsis) recommendations have been, and will be, followed where relevant.^18^ Construction of the BPGs will be guided by the refined version of the Appraisal of Guidelines for Research & Evaluation (AGREE II) instrument and reported following the Good Reporting of A Mixed Methods Study (GRAMMS).^19,20^

The BPGs will be produced using a mixed methods approach, allowing for the integration of the three pillars of evidence-based medicine: best available evidence, patient perspectives, and clinical reasoning (Figure 1).^21,22^ The overall design is a two-phase segregated, convergent design,^23^ with four components (three pillars plus focus groups), consistent with our previous approach.^15,16^ Phase 1 will focus on generating valid quantitative and qualitative data addressing the three pillars of evidence-based medicine. Best evidence will be synthesised through systematic reviews with meta-analysis of high-quality studies, while patient values will be elicited through in-depth, semi-structured interviews with people recovering from, or who have recovered from, the condition, followed by thematic analysis. Clinical expertise will be synthesised through in-depth, semi-structured interviews with multidisciplinary experts in the management of the condition, followed by thematic and content analysis. Finally, focus groups or stakeholder engagement events will be conducted with experts to review the format and structure of the final BPG.

**Figure 1.**
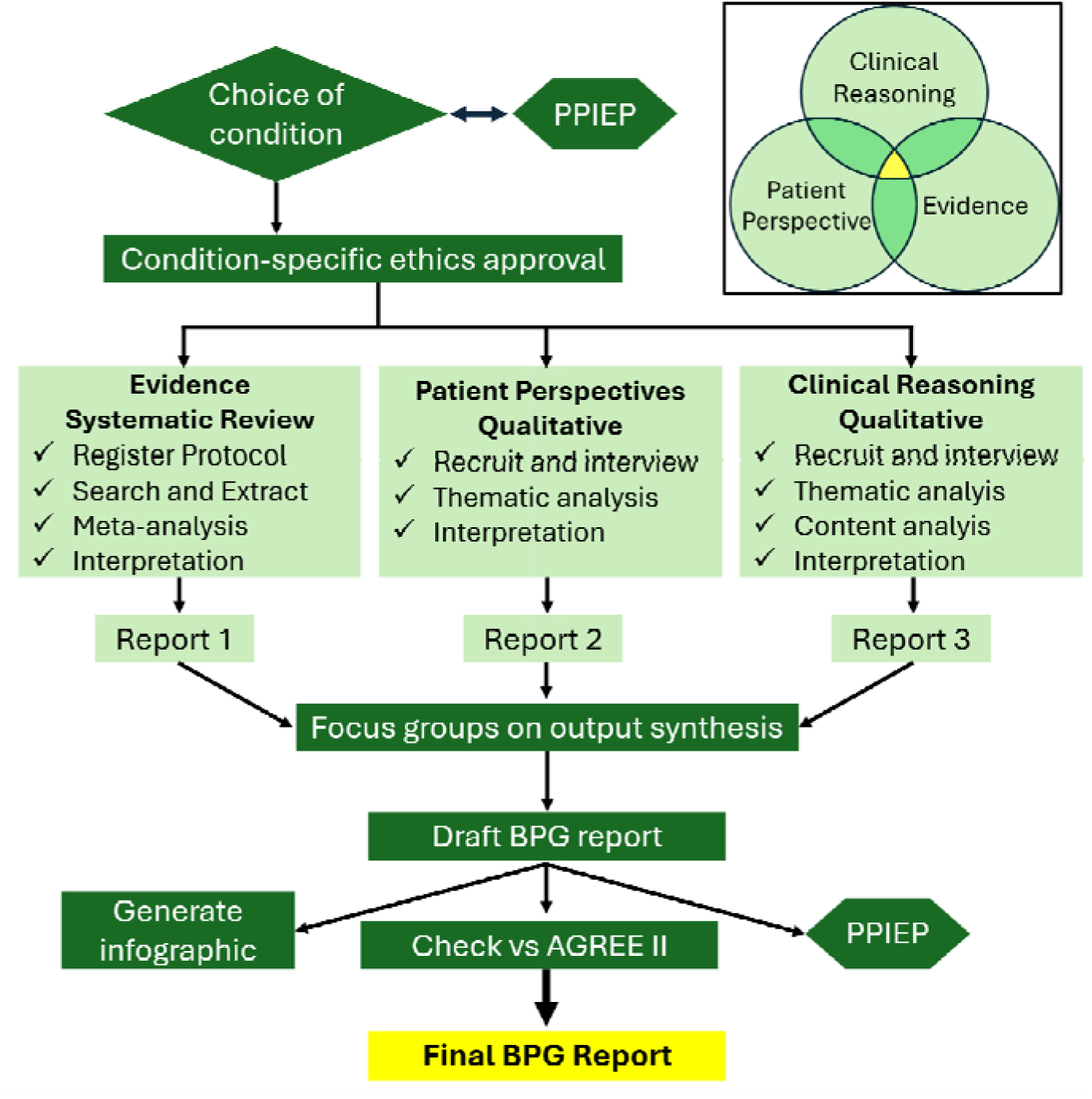
Integration of the three pillars of evidence-based medicine for the construction of a Best Practice Guide.

Although guided by predefined protocols, data collection does allow early systematic review findings to shape the later stages of the qualitative studies. Integration mainly occurs at the analysis stage, where the different data streams are brought through integrative analysis and focus groups in phase two. This phase is intended to deepen interpretation and reduce threats to validity by ensuring data streams remain aligned in their messages and outputs.^24^

The methods used to produce each BPG are summarised in **Error! Reference source not found.**, and derive from our recent published BPGs– particularly those for plantar heel pain and patellofemoral pain.^15,16^ What to include in the BPG will come predominantly from the systematic review and meta-analysis, while when and how to combine and apply interventions will mainly be derived from the interviews. That is, specific interventions are therefore mainly determined by the review, whereas service delivery, clinical reasoning, and confirmation of patient acceptability will be informed by the qualitative studies.

### Public and Patient Involvement, Engagement and Participation (PPIEP)

PPIEP is the active involvement of patients, carers, stakeholders and the public in all aspects of research, from the design of studies to the dissemination of findings, and has been shown to have meaningful impact on guideline development.^25^ PPIEP has been, and will continue to be, central to the research design, conduct, and dissemination of the BPG.

Prior to the start of data collection for each condition, at least two meetings with patients and clinicians involved with each condition will be conducted. These will serve to guide selection of relevant outcomes for the systematic review and creation of the topic guides for the qualitative studies. Throughout the project, further focus groups with experts will be conducted to present current results and discuss data interpretation and next steps.

Within each meeting, we aim to select participants that are diverse in age, gender, ethnicity, and professional background. The activities are fully costed, including for attendance at face-to-face meetings. We have allowed ∼£27.50 per participant as compensation for participant time, to encourage recruitment but not be financially coercive, based on national guidelines.^26^ Additionally, we will invite at least one patient per condition to join the study as a co-author. Patient involvement at this stage is essential to clarify the clinical significance of the findings, enhance the quality of reporting, and ensure that the research question addresses issues that are meaningful to patients.

### Selection of conditions

The choice of topics for BPG formulation was made at an early study design stage. Both the criteria for selection, and proposed choice of conditions, were reviewed in the first PPIEP meeting, which generated additional criteria. These criteria were divided into the details of the condition (clearly defined condition, strong recent evidence, high incidence of problem, variable treatment and support of PPIEP group) and availability of current guidelines (clear guidelines, well-defined for physiotherapy, patient voice included and within scope of practice). Each criterion was graded on a 3-point scale, leading to the selection of Rotator Cuff Tendinopathy, Lateral Elbow Tendinopathy, Acute Lateral Ankle Sprain and Lumbar Radiculopathy as the initial topics for BPG formulation. Gluteal tendinopathy had already been started as part of a related PhD. Plantar heel pain, and patellofemoral pain were in late stages of completion. Further PPIEP meetings will be held to select the final conditions.

### Ethics and Governance

Ethical approval for the qualitative studies will be gained from Queen Mary University of London’s Research Ethics Committee for each condition separately. Ethics will also be obtained from the NHS to facilitate recruitment of patients with the selected conditions. Ethics is important in clinical guidelines research because of the meaningful impact on the care that patients receive, therefore ensuring research integrity and bias minimisation is essential.

### Systematic review

#### Overview

The systematic review methods may be refined as we progress through the six conditions, to reflect changing software support and the particular methodological challenges associated with different conditions. These particularities will be detailed in the individual PROSPERO registrations.

The review will be aligned with Cochrane and Joanna Briggs Institute (JBI) methodological guidance for reviews of interventions by a team with extensive experience and world-leading methodological expertise in systematic reviews. The protocol for the systematic review elements has been registered on PROSPERO (Plantar heel: CRD42018102227, Patellofemoral: CRD42019152252, Gluteal: CRD4201914023, Shoulder: CRD42024584126, Ankle: CRD420251244514, Elbow: CRD42024584431 Lumbar: CRD420251246729 and will adhere to the Preferred Reporting Items for Systematic Reviews and Meta-Analyses (PRISMA) statement, including the extensions for network meta-analysis, complex intervention, and search reporting.^27–29^ The reviews will follow a dual framework: the primary comparative tool will be a predictive meta-analysis comparing a benchmark intervention (or minimal intervention) with the changes resulting from active treatments, complemented with a network meta-analyses.

#### Information sources

PubMed, Embase, Web of Science, CINAHL, and SPORTDiscus will be searched from inception without language restrictions or filters for eligible randomised controlled trials (RCTs). Reports in languages other than English will be translated using software or by colleagues/collaborators of the research team.

Search strategy will be created based on other systematic reviews, with terms used to identify the population and correct study design. Interventions, Comparators and Outcomes will not be specified as all interventions and comparators will be eligible (except exclusively surgical techniques comparisons) and tools for measuring outcomes are too varied and often omitted on study tile and abstract. Forward and backward citation tracking of included studies will be conducted through the Scopus database and by screening publications from known researchers in the field.

#### Study selection and study quality

RCTs containing at least one arm of conservative treatment for the management of the condition and measurement of at least one of the selected outcomes will be considered. Studies only comparing different types of surgical treatment or that do not report any of the outcomes will be excluded. Specific inclusion and exclusion criteria regarding population and minimum follow-up will be defined for each condition.

Records will be exported to Covidence (Veritas Health Innovation, Melbourne, Australia), duplicates removed and non-RCTs automatically classified and excluded.^30^ Titles and abstracts will be independently assessed by pairs of reviewers with full texts being assessed by one reviewer with all exclusions being confirmed by a second reviewer. Disagreements will be resolved by discussion.

An additional inclusion criterion for these systematic reviews was for the report to be of high methodological quality and low risk of bias,^15,16^ which will be evaluated using the PEDro scale.^31^ An extra item evaluating conflict of interest by the authors or funders will be added to these evaluations, with studies that report relevant conflicts or do not mention conflicts not being awarded a point. Thus, reports must score at least eight points in the PEDro scale and six points in the items relevant to risk of bias found in the PEDro scale.^32^ These thresholds may be adjusted for different conditions. Where necessary, the quality and risk of bias criterion may be waived to include adequate numbers of studies with minimal interventions. A single reviewer will evaluate the reports, with those on the threshold of inclusion (i.e., 7 and 4 points) being confirmed by a second reviewer. The results of searching and selection, as well as reasons for exclusion at full text screening, will be presented in a PRISMA flow diagram.

#### Data extraction

Data will be initially extracted using the Elicit tool (Elicit Research PBC, Covina, USA), where the study texts will be uploaded and carefully developed prompts given for the extraction of relevant data (Table 1). All data extracted by the tool will be manually confirmed by one investigator and completed when missing. Published study protocols, supplementary material, and study registration will also be manually checked to complete or correct extracted data. Where data are missing, or not in a usable format, we will attempt to contact the corresponding author of the study up to two times, and will otherwise impute where possible according to the Cochrane Handbook for Systematic Reviews of Interventions.^33^ The data extracted will include participant demographics (e.g., age, symptom duration, symptom severity, and BMI), study design (e.g., outcomes, time points), intervention characteristics, and group mean and distribution values. Symptom severity will be measured using the most common tool used for assessing the condition, when possible, with less common tools used when necessary. Outcomes extracted and relevant time points will be selected for each condition through PPIEP sessions. Interventions will be divided into their component treatments, with data regarding duration, frequency, location, adherence, and fidelity extracted for each treatment.

**Table 1.** Data categories extracted and prompts given to the Elicit tool for structured automatic data

| **Data category** | **Prompt given** |
| --- | --- |
| **Aims and Purposes** | What are the study's aims or purposes? |
| **Additional Data** | Does the study indicate that there have been other articles published using data from the same study? Does the study have a protocol paper published? Does the study have any supplementary material? What does it contain? |
| **Condition Description** | How does the study call the condition investigated? |
| **Age/BMI/Symptom severity** | What is the average and distribution of age/BMI/symptoms of all the participants that were evaluated at baseline? Pooled average of all groups if the data is presented separately. Prioritize data reported on the table. |
| **Sex** | What is the percentage of females that were evaluated at baseline? |
| **Physical Activity** | What is the level of physical activity from the participants included in the study? Do not consider any activity started after the start of the study. Choose one of these options: 1 = Athlete; 2 = Physically active; 3 = Sedentary; 4 = Not reported |
| **Interventions** | List the interventions that participants received. |
| **Intervention details** | Identify which component treatments were given to the patients in each Intervention. As an example, if the patients received only electrical therapy, there would be only one treatment. However, if the patients also received exercise, then there would be two treatments.<br>Treatments that only some participants in the group received do not count. This usually would be painkillers and would be reported that they had the option of taking them.<br>Choose one of possible classifications of elements within an intervention (*condition-based list given*)<br>Find the following information about each component treatment:<br>• Name of the group - Name used by the authors to refer to the group<br>• Duration - For how many weeks did the participants receive this component treatment? If reported in months or sessions, convert to weeks. If unclear report it as "unclear" and make sure to justify it in the explanation.<br>• Frequency - How often did participants receive this component treatment per week? If reported as number of sessions, convert to times per week. Pay particular attention when an element is given multiple times a day (as opposed to weeks). In these cases, frequency will be the number of times a day multiplied by 7.<br>• Development - Does the study describe how the component treatment was developed? Development will usually involve a pilot study or a previous study. If yes, give brief description. If no, report it as "not reported".<br>• Adherence - Did the participants actually follow the component treatment? Write any information about it. If not reported, report it as "not reported"<br>• Fidelity - Did the participants follow the component treatment correctly? Injection and passive treatments (electrotherapy, laser, manual therapy, etc) will automatically be yes. If all exercises in the program were supervised, this will also be yes (reported as "supervised"). When exercises or other element was done at home, then the authors need to describe how fidelity was assessed. If not reported, report it as "not reported" |
| **Outcomes** | If one or more outcomes were defined as the main/primary outcome by the authors write its name before anything else as "Primary outcome = names of outcome" or as "Primary outcome = not reported"<br>Include all measured outcomes. Find the scale in which each outcome was measured in (min-max) and if positive values were good or bad. |
| **Timepoints** | Give the timepoints in which participants were evaluated. Report them in weeks.<br>If the timepoint was measured from the end of the treatment (as opposed to from baseline), report it.<br>Reorganize each time point into four categories (*may vary with condition*):<br>• Baseline = Any measurement made before the start of treatment<br>• Short-term = Between one week and 3 months included<br>• Medium-term = Between 3 months (excluded) and 12 months (excluded)<br>• Long-term = 12 months or longer (if only longer is available) |
| **Results** | For each intervention/outcome/timepoint report the average value (mean or median) and distribution values (standard deviation, standard error, range, min-max, interquartile range or 95% Confidence interval). If one of them is not reported place "NR" instead of the value. Baseline results may be on another table (typically participant characteristics table). Prioritize data reported on the tables.<br>For each intervention/outcome/timepoint, identify how many participants were evaluated in each intervention and in each timepoint. If the number is not reported for each timepoint, use the baseline number. This is likely to be in the flow chart diagram/Consort diagram. |

#### Intervention and treatment classification

Initially, we will identify whether the focus of the intervention is clear or not. A clear focus will be identified through the presence of the treatment name in the report title, aims or name given to intervention group. Those without this will be considered to have an “unclear focus”. Subsequently, a set of rules will be in place to classify interventions into different treatment foci to pool for statistical comparison (Table 2). Treatments given within an intervention that are not considered the focus will be classified as adjuncts.

**Table 2.** Categories of intervention focus and description of criteria to be included in each one. Divided into cases where the focus is clear or unclear.

| **Focus (determined by presence in study title, aim or group name)** | **Description** |
| --- | --- |
| **Clear focus: Treatment was mentioned in the study title, study aim or group name** |  |
| **Complex Intervention** | Interventions with three or four plausible treatment types will be classified as complex AND will maintain their individual foci |
| **Impairment-based Multimodal physiotherapy** | When multiple treatment types are available and specific ones are chosen based on individual patient impairment will be classified as such and not have any individual focus |
| **Highly-Complicated Multi-treatment Interventions** | Interventions with five or more treatment types will be classified as such and not have any individual focus |
| **Unclear focus: Often referred to only as “physiotherapy” or “control”** |  |
| **Multi-focus** | If there are less than three treatment types, each treatment will be classified as a focus |
| **Complex Intervention** | Interventions with three or four plausible treatments types |
| **Impairment-based Multimodal physiotherapy** | When multiple treatment types are available and specific ones are chosen based on individual patient impairment |
| **Highly-Complicated Multi-treatment Interventions** | Five or more total treatment types |
| **Unclear Complex Intervention** | Unclear focus and no well-reported treatment types |

Besides the classification into treatment foci, interventions with clear focus will additionally be classified as “Complex Intervention” if they include three or four plausible treatments. Interventions with five or more treatments will be classified as “Highly-Complicated Multi-treatment Interventions” and will not enter the analysis as they are too varied to permit attribution of observed effects to individual components.

#### Data synthesis

Treatments will be classified and pooled into both broad (e.g., exercise, injection, electrotherapy) and more specific categories (e.g., strengthening or range of motion, steroid or hyaluronic acid injection) enabling different levels of aggregate synthesis to better establish the most effective approaches. Treatments will be combined based on clinical interest, plausibility, and underlying rationale. For example, within the Rotator Cuff Tendinopathy review, stability, proprioception and motor control exercises will be grouped into Movement Pattern Retraining, which will be further grouped (with strengthening and range of motion) into Exercise (Appendix 1).

The predicted distributions expected in future studies for each treatment will be obtained for each outcome and time point using Bayesian random-effects meta-analyses. These will then be used to estimate the probability that each active treatment would be superior to a benchmark in a future trial, using posterior predictive simulations of expected distributions (m = 10,000 iterations). Benchmarks used will be the expected distributions of minimal intervention (wait-and-see or sham), the minimal clinically important change (MCIC) or the expected distribution of another active and common intervention, when the former is not available.

Network meta-analyses will be conducted for each outcome at each time point to enable comparison across treatments by integrating head-to-head evidence and indirect comparison. Network meta-analyses will be conducted for each outcome at each time point to enable comparison across treatments by integrating head-to-head evidence and indirect comparison. Sensitivity analyses will be conducted according to data availability and clinical interest. These may include: (1) population subsets, (2) a higher level of treatment resolution (e.g., “PRP” instead of “Injection”, Appendix 1), or (3) including or excluding studies with different PEDro scores or risk of bias.

#### Certainty of Evidence (GRADE)

Grading of Recommendations, Assessment, Development and Evaluation (GRADE) will be used to determine the certainty of evidence for each analysis framework. For the benchmarked analyses, certainty will be assessed for each treatment-versus-benchmark comparison at each outcome and time point. For the network meta-analyses, certainty will be assessed across the network estimates for each outcome and time point. Certainty of evidence will start as high and be adjusted according to specific objective criteria, which will be evaluated by one author. These will guide final recommendations based on evidence synthesis.

### Risk of bias

Risk of bias will be further assessed using the Cochrane Risk of Bias 2 tool (RoB2). The Elicit tool (see below for details) will be used to extract relevant excerpts from the text using the signalling questions and elaboration provided. One investigator will apply the RoB2 tool to all studies, classifying each domain and the overall studies as high, some concern or low risk of bias. Certainty will be downgraded one level if ≥50% and <75% of pooled data comes from studies with high risk of bias or some concerns and downgraded two levels if ≥75% of pooled data comes from studies with high risk of bias or some concerns. A second investigator will also apply the tool to a sample of studies to confirm accuracy.

### Publication bias

Publication bias will be assessed by sampling trial registrations from the International Clinical Trials Registry Platform across three representative periods. For each period, a random sample of trials will be chosen, and a single researcher will screed for intervention focus and publication status. We will consider suspected publication bias when unpublished trials outnumbered published trials for a given treatment category, which will result in downgrading certainty of evidence by one level.

### Imprecision, inconsistency and indirectness

Imprecision will be judged based on the magnitude of the posterior probability that a treatment would be superior or inferior to the benchmark of minimal intervention. Downgrading by one level will be applied to probabilities classified as low (0.60-0.75) or very low (0.50-0.60) certainty, as they will be considered to reflect serious imprecision. Inconsistency will be determined based on the posterior median between-study standard deviation, with thresholds being selected according to the outcome chosen for the condition. Indirectness will not be used to downgrade certainty of evidence as the included populations, interventions, and outcomes will be directly relevant to the review question. For the network meta-analysis, imprecision will be judged primarily by the stability of treatment rankings across the posterior distribution, with greater uncertainty where rankings vary substantially. This will be interpreted alongside the width of the 95% credible intervals and their position relative to the null effect and, where available, thresholds for clinical importance. Inconsistency will be judged from incoherence between direct and indirect estimates and the magnitude of between-study heterogeneity.

### Patient perspectives

#### Overview

Patient perspectives will be elicited through semi-structured, in-depth interviews. Analysis and reporting will be guided by the TACT framework and the COREQ checklist. The analyses will be positivist for expert clinical reasoning and interpretivist for patient perspectives, both analysed using the Framework approach.^34^ Reflexivity statements for the researchers will be provided as we acknowledge that researcher background may influence data collection and interpretation.

#### Patient recruitment

Recruitment will be facilitated via advertisements in private clinics, NHS clinics, social media advertising and approach to patient support groups as available. Possible participants will be approached in person or through email and, if they are happy to take part, will answer initial questions to confirm eligibility. Those eligible will be asked to complete the consent form after reading the patient information sheet and asking any questions needed. Once consent has been obtained, the interview will be schedule at the patient and researchers’ earliest convenience.

We will adopt the approach described by Malterud (2016) that prioritises information power rather than data saturation.^35^ Our experience is consistent with the literature that reaching data sufficiency and high information power requires at least 10 and up to 20 participants per group of experts and similar numbers for patients. Further, we will aim to include a representative sample by recruiting patients with varied characteristics across nationalities, age, sex, care pathways and condition presentation. We will allow a monetary compensation for interviewee time, to encourage recruitment but not be coercive, based on NIHR guidance, and our own Centre for Public Engagement (approximately £25).

To ensure transferability of findings related to patients’ lived experience, we will use a purposive sampling frame to include age, sex, ethnicity, country, duration of symptoms and stage of condition. Each participant will have to meet the diagnostic criteria for the condition of interest, have had experience of treatment and speak adequate English or a language in which the interview team is fluent.

#### Interview process

One researcher will conduct each interview online using Microsoft Teams, lasting ∼60-90 minutes. An interview topic guide will be used, which will be constructed based on a preliminary literature search, discussions within the research team, with PPIEP group members and guided by emergent concepts from pilot interviews. Questions will explore interviewees’ experience of the condition and their management, including perceptions of the interventions, information, and education they received to date, and their interactions with clinicians. Answers for the Patient Acceptable Symptom State (PASS), Single Assessment Numeric Evaluation (SANE) and Global Rating of Change (GROC) will be recorded at the start of the interview to quantify patient status. An online graphic tool will be used to help the interviewer and the patient to visualize the timeline of recovery, with boxes, arrows and lines representing the onset events, assessments, treatments and perception of recovery at different points in the trajectory (Figure 2). This tool is not restrictive as it does not follow a strict timeline and is used later in the interview. The final resulting graphic will be sent to patients for confirmation.

**Figure 2.**
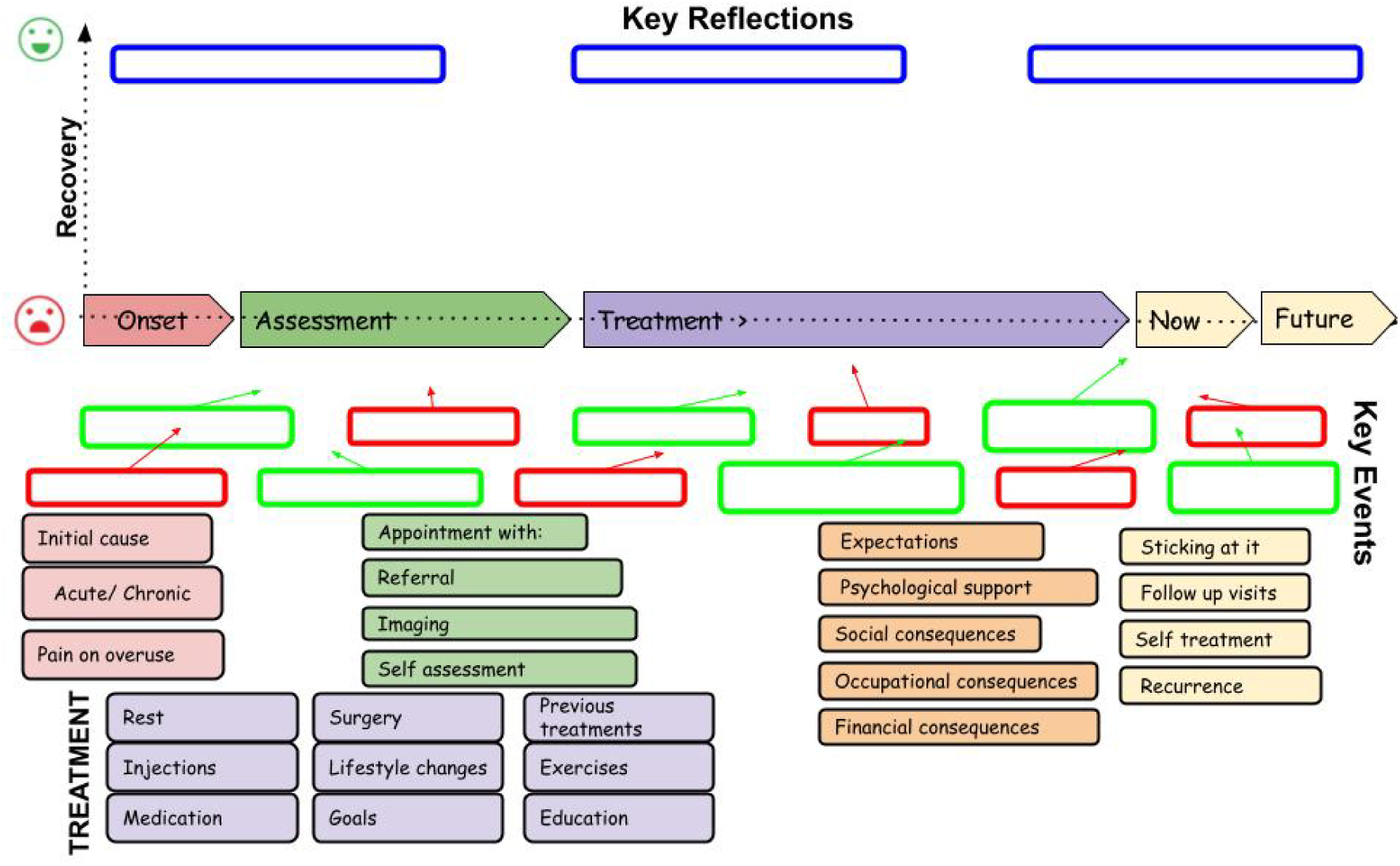
Example of blank diagram tool that will be used to help elicit and organise patients’ experience.

#### Data analysis

Analysis will be conducted concomitantly with data collection. Recording of interviews will be uploaded into notta.ai (Notta Inc., Tokyo, Japan) and manually checked by one researcher to generate an accurate transcript, which will then be uploaded to Atlas.ai (ATLAS.ti Scientific Software Development GmbH, Berlin, Germany) for initial coding.

Data will be analysed thematically using a pragmatic framework approach, which may differ according to the condition.^34^ Using a mixed inductive-deductive approach, initial codes will be generated based on the data while acknowledging that some themes will be similar to those pre-determined in the topic guide and seen in similar studies. Codes will be combined or added as interviews are analysed according to code similarity or lack of appropriate code. Once all transcripts have been coded, data will be exported into Excel, where initial themes, subthemes and findings will be identified. Each subtheme will be summarised and representative quotes identified for each finding. Multiple iterations of combining or separating themes, subthemes and findings will be allowed until the researchers agree on the final results. When disagreements are present, researchers will discuss until consensus is reached.

### Clinical Reasoning

#### Overview

Clinical reasoning will be elicited using a similar approach as patient perspectives. Differences will be regarding participant inclusion criteria, the areas of questioning, the graphical tool used to assist in clinical reasoning and the addition of a content analysis of individual treatment options.

#### Recruitment of clinical experts

International experts are defined, as per previous methods,^15,36^ as having a minimum of 5 years of experience in a setting and specialty in which they regularly encounter significant numbers of people with the condition of interest and are actively involved in relevant research or evidence translation. Experts will be identified by the authors through recommendations of researchers that publish in the relevant field and snowball sampling. Experts will also be directly invited to participate via an approach email, with the text having been ethically approved. To ensure transferability of clinical reasoning findings, we will use a purposive sampling frame for world experts to include age, sex, primary occupational sector, country and profession.

#### Interview process

For international experts, questions will explore interviewees’ background, clinical reasoning when managing the condition, perceptions of the evidence and any gaps in the published literature. An online graphic tool containing descriptors of possible interventions will be presented at mid-interview as a stimulus to discussion. This tool requires experts to place different treatment approaches (non-exhaustive list presented in the tool) across an example treatment timeline (Figure 3). Experts will be invited to complete the tool on their own device, while describing their reasoning for choosing each treatment application.

**Figure 3.**
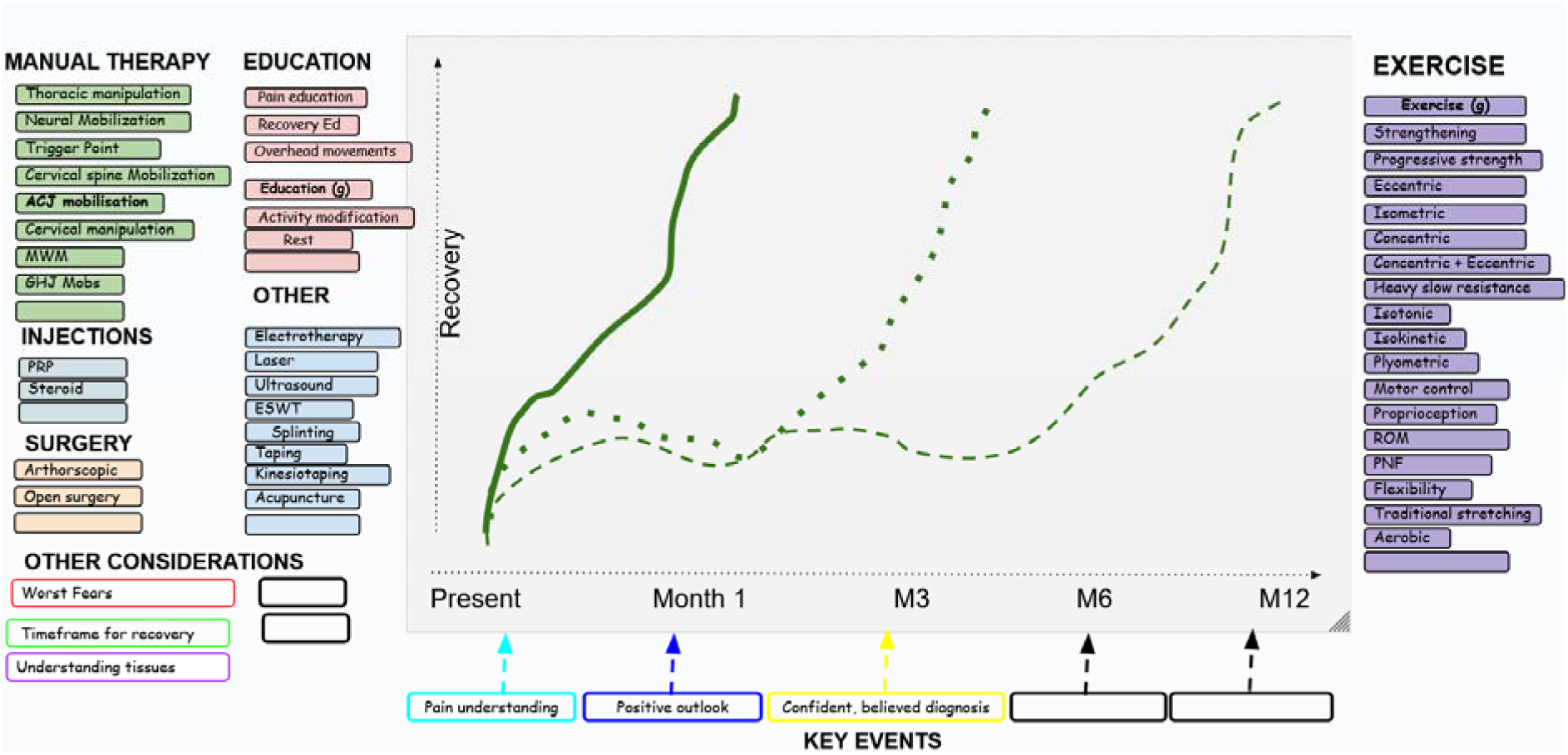
Example of blank diagram tool that will be used to help elicit clinician’s reasoning for choosing treatments.

#### Data analysis

In addition to the thematic analysis described above, we will also conduct a content analysis to identify experts’ beliefs on individual treatments, and combinations of treatments into interventions. Items identified will be the mechanism of action, how they identify if the treatment is successful, their thoughts on the current level of evidence and method of optimal delivery, including when to apply, how to progress, which treatments can be used in combination, and which cautions should be taken. Based on experts reasoning (data volume and consistency of perspective), each treatment will receive a final recommendation.

### Combination of findings and focus groups

In addition to the mid-project focus groups (see PPIEP), one last set of focus groups will be conducted once all data has been analysed. These will be conducted with a sample of experts interviewed previously and will aim to ensure the clinical reasoning closely maps to the interventions with proof of efficacy. This process has been used successfully by the research team previously and is congruent with recent developments in internationally recognised guideline development methodology, particularly the inclusion of qualitative evidence in guidelines.^37^

Additionally, focus groups will serve to confirm the scoring system used to give a final recommendation to each treatment type, combining the findings from the three data streams. These recommendations will have a numeric value, which will be further classified into “not recommended”, “low recommendation strength”, “moderate recommendation strength” and “high recommendation strength”. Initial proposed scoring system will be as follows, with focus group guiding weight and inclusion/exclusion of each item.

- Systematic reviews will result in a maximum of 18 points (6 maximum points for each timepoint) originating from:

`o` Probability of being superior to the benchmark (minimal intervention, MCIC or other common intervention) – Maximum 4 points
`o` Effect size of the change from baseline – Maximum 2 points
`o` Certainty of Evidence (GRADE) – Maximum 0 points with one point being subtracted from each of the five items
- Clinical reasoning will result in a maximum of 6 points according to the level of recommendation. Recommendations against the treatment type will result in subtraction of a maximum of 6 points.
- Patient perspectives will result in a maximum of 3 points according to patients’ support for the treatment. Points will be subtracted in cases where patients are against the treatment.

### Dissemination

BPGs will be widely disseminated and implemented into practice through multiple approaches. The BPGs will initially be disseminated through the traditional methods (i.e., publications and presentations). Importantly, a learning package will be developed and made freely available via an online platform. Further, dissemination routes that involve partnerships with clinical and academic institutions will be developed and employed to assist in the dissemination.

## RESULTS

### Outputs

The primary research outputs will be four reports per condition, submitted for peer review, detailing the findings of the systematic review, the two qualitative studies and the main BPG.

### Dissemination

The BPG will be translated into clinical practice using a range of educational materials via a freely available learning package (Figure). These will be hosted on a website or on an online learning environment. Two main educational pathways are planned: (1) an educational resource package, containing learning of content generated by each BPG and its component studies and (2) a moderated learning community, where example patient cases will be used to evaluate learning through case description and peer feedback. Each of these pathways will be properly certified within each country’s higher education system.

**Figure 4:**
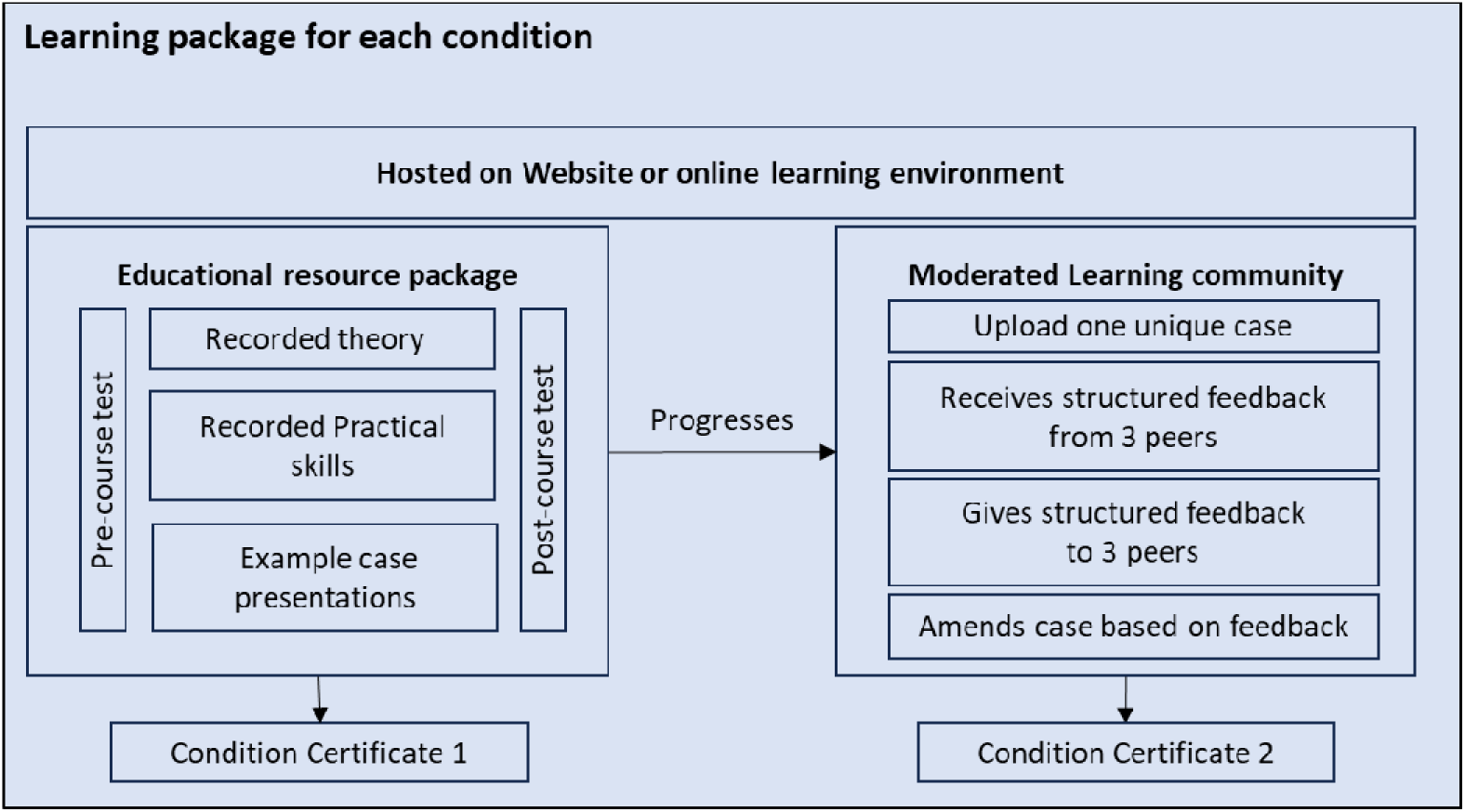
The components of the learning package for each condition are outlined, alongside the requirements for participating student or qualified physiotherapists to realise credit.

**Figure 5:**
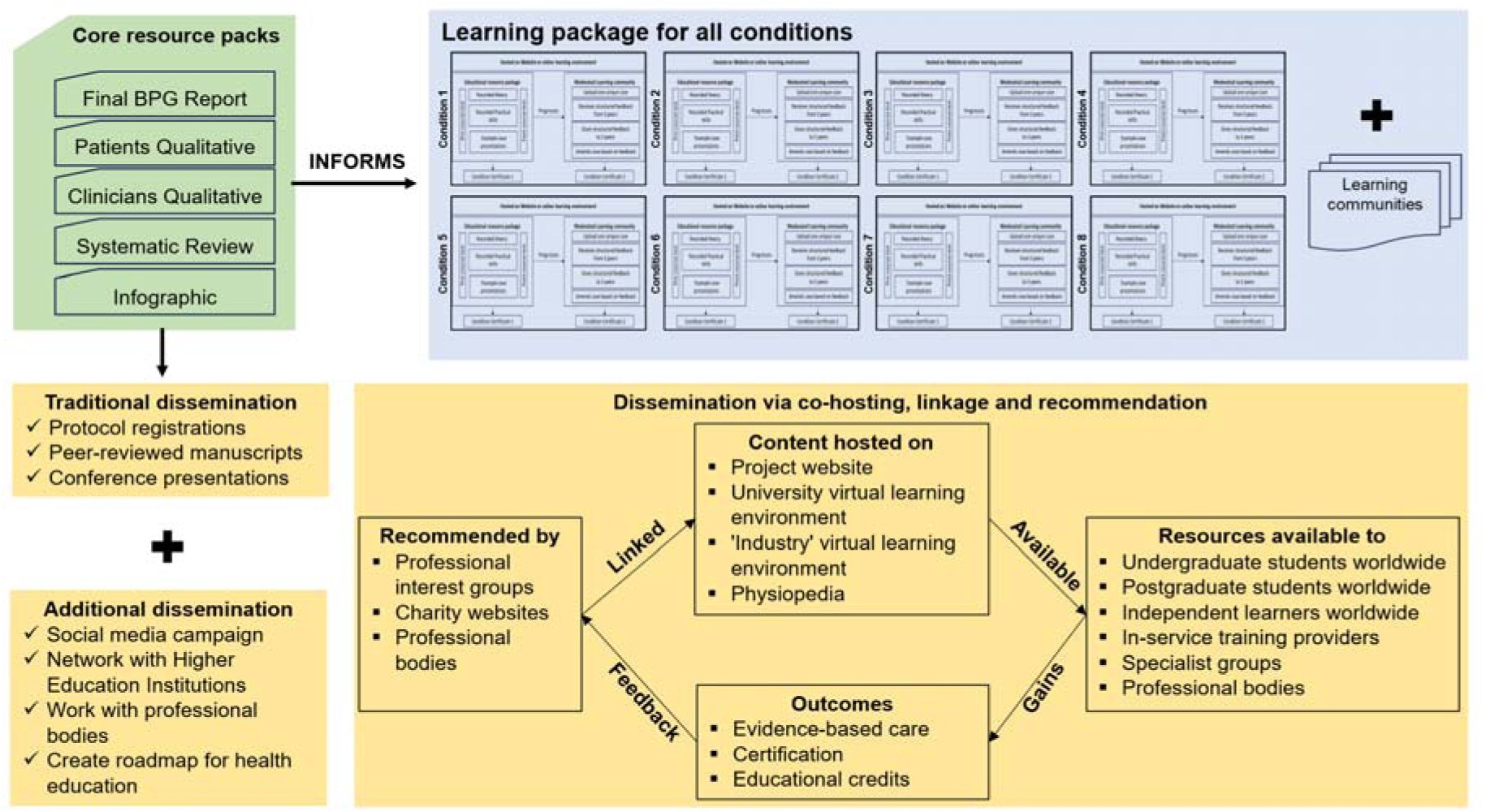
Traditional dissemination routes are indicated alongside the innovative dissemination package of educational content. The illustration shows how the content can be linked to, hosted, and utilised by recipients. The outcomes, including patient care advances are also indicated.

A second layer of dissemination will include networking with a range of key stakeholders who deliver education, set policy and can recommend the BPG to relevant practitioners, as outlined in Figure. These will be conducted within the United Kingdom but also internationally in countries such as Australia, Brazil, Greece, Indonesia and Pakistan, aiming to reach geographical and contextual diversity.

## DISCUSSION

### Guideline development

Clinical guidelines are defined as ‘systematically developed statements to assist practitioner and patient decisions about appropriate health care for specific clinical circumstances’.^38^ Several approaches to the development of clinical guidelines are present in the academic and grey literature. Many are produced by research groups, professional bodies or institutions without a transparent description of the methods used to develop the recommendations, making it difficult to assess methodological quality or potential biases. This can limit credibility and, consequently, the adoption of these guidelines. Therefore, structured instruments such as the AGREE II appraisal tool have been developed to assess and improve the process of guideline development and reporting.^20^

Other approaches such as the National Institute for Health and Care Excellence (NICE) guidelines have emphasized a rigorous process and are widely regarded as a benchmark for evidence-based recommendations in the UK.^39^ Although these guidelines are methodologically rigorous, involving systematic review of evidence, economic evaluation and stakeholder consultation, they are often limited due to a lack of clear, clinical applicability at the individual patient level and typically hampered by the lack of adequately powered cost-effectiveness studies in the musculoskeletal field. Critically, they lack the detail of empirically elicited clinical reasoning and patient perspectives.

Another commonly used method to generate recommendations for clinical practice is the Delphi technique, a method that uses multiple rounds of questionnaires with feedback to achieve consensus among experts.^40^ It is a useful method where data is sparse but is almost entirely dependent on expert opinion, lacks direct incorporation of the published evidence, and typically neglects patient’s perspectives - an important domain in the AGREE II tool. Results may be influenced by panel composition, power dynamics within the participants and inherent expert bias towards a given approach. Therefore, Delphi-derived recommendations may reflect prevailing beliefs from a limited group rather than robust evidence.

### Mixed-methods

The advantages of integrating qualitative and quantitative evidence sources in healthcare research are widely acknowledged,^15,16,24,41^ particularly in relation to complex interventions,^42^ with qualitative evidence syntheses being increasingly used by guideline developers.^39,43,44^ By integrating primary qualitative evidence with systematic review findings, we will base recommendations on effectiveness of interventions on the highest-quality RCT evidence and will importantly be able to contextualise recommendations based on patients’ and experts’ experiences. Whilst there is some existing published qualitative evidence on the conditions of interest, conducting novel qualitative inquiry will allow us to focus on aspects specific to supporting BPG recommendations, instead of more generalised experiences and perceptions of conditions and their management. Ultimately, the systematic review will tell us what works, while patients will tell us what and how those treatments will be used and clinicians will tell us when, how and how much each treatment should be given. Integration in mixed methods approaches is often superficially reported. To this end, and drawing on contemporary discourse,^45^ we will adopt a full integration approach,^46^ intentionally integrating data sources throughout the project (see Figure 1), so that regular ‘conversations’ between data sets generated from multiple methods inform the final guidance.^45^

### Planned updates

The BPG’s will require updating to reflect advances in knowledge and the increasing evidence base. In the case where a novel or markedly improved treatment results in a step-change in outcomes for a given condition or sub-group, then the guidelines would require a complete overhaul. This would apply particularly to the systematic review although some updating of the clinical reasoning would also be required. More likely, for all guidelines, is that a steady accumulation of high-quality evidence will enable stronger recommendations to be made at a higher degree of resolution. In other words, the detailed prescription for particular treatments, for subgroups of patients, stages of presentation, and for particular contexts may be revealed through higher power and better certainty from systematic reviews. Further, more outcomes will become ripe for evaluation, such as cost-effectiveness or biomarkers. To an extent, this has been our experience with revising the patellofemoral pain BPG.^16,47^ Further, methodological innovations may enable guidelines to be made available in new ways – for example as clinical decision support tools embedded in usual-care clinical systems, thus addressing one of the current barriers to their adoption.^48^ It is envisaged that AI tools will increasingly be accurate enough to update reviews, albeit keeping the ‘human-in-the-loop in order to reinterpret the evidence.^18^

### Developing role of AI

Artificial intelligence (AI) is a promising addition to systematic review methodology when applied transparently and with appropriate safeguards, offering meaningful efficiencies.^49^ In this programme, we will build on our growing experience using AI-assisted approaches for literature searching, risk-of-bias assessment, and data extraction. Such approaches are likely to be particularly valuable for living systematic reviews, given the recognised delay between searching and publication, the limited frequency of updating, and the challenge of sustaining review maintenance after the initial funded phase.^50^ Recent evidence indicates that AI and semiautomated tools are currently used most often for data extraction or collection and risk-of-bias assessment, whereas their use for publication updating remains limited.^50^ We will therefore aim to establish simplified post-project update processes, supported where appropriate by automated and semiautomated tools and combined with targeted human oversight, to enable efficient incorporation of smaller volumes of newly emerging evidence with lower ongoing resource requirements.

## PPIEP

Integrating PPIEP strengthens the overall design, relevance and anticipated implementation of the BPGs. Early and ongoing involvement of people with lived experiences of common MSK conditions ensures that the planned methods, outcomes and dissemination strategies are aligned with the various stakeholder’s priorities, improve clarity and accessibility and address the potential barriers to engagement and equity from the onset.^51,52^ The NIHR guidance emphasises that including lived experiences from the onset increases the reach, relevance and impact of health research, and is an essential component of best practice in protocol development. Integrating PPIEP at the protocol stage, therefore enhances transparency, quality improvements in MSK care with patients and for patients and likelihood that the BPGs will be acceptable and useable in the real-world settings.^53^ This supports a co-produced approach that is increasingly recognised as essential for producing actionable, patient centred MSK guidelines.

### Dissemination and implementation

Clinical guidelines frequently fail to translate into routine practice because their recommendations do not align with the contextual, organisational and behavioural realities faced by clinicians.^54^ Further, clear, consistent condition definitions are essential for developing clinical practice guidelines^55^ as they eliminate terminological ambiguity, enable valid comparison and synthesis of evidence and support uniform interpretation across stakeholders.^20^ Even when guidelines are disseminated effectively, uptake falters without organisational structures such as leadership buy in, decision support systems, and protected staff training time to enable clinicians to familiarise themselves with the guidance.^56^ Previous research reports that lack of knowledge, insufficient training and low clinician confidence in using the guidance were key reasons clinicians did not use guidelines despite availability.^54,56,57^ The guidelines themselves can lead to implementation failure. Limited usability and complex guideline design, poorly structured, overly technical, or excessively lengthy guidelines can impede implementation because clinicians cannot easily extract actionable recommendations.^57^ Co-creation of clinical guidelines with stakeholders is crucial for improved dissemination and implementation, a key component of the BPG protocol.^15,58^ These issues are addressed in each condition-specific BPG.

The Best Practice Guide (BPG) model directly addresses the recognised barriers to dissemination and implementation. Definitional ambiguity is resolved by establishing a consistent, evidence!ZIderived condition definition, enabling uniform interpretation across research, education and clinical settings. The mixed-methods structure explicitly incorporates real-world clinical reasoning and patient values, ensuring that recommendations reflect the contextual, organisational and behavioural realities clinicians face rather than idealised trial conditions. Co-production with clinicians and patients throughout development further enhances relevance, addressing barriers such as lack of stakeholder involvement and low perceived feasibility.^59^ Finally, each BPG will be paired with a condition specific educational package that supports clinicians’ knowledge uptake, practical skills and confidence directly mitigating implementation challenges arising from limited awareness, insufficient training and uncertainty about how to apply guidance in practice.

### The challenge of measuring impact on patient outcomes

A further challenge is that any effect of a best practice guide on patient outcomes is inherently difficult to attribute directly. Clinical guidelines do not act on patients in isolation; rather, their influence is mediated through clinician awareness, adoption, contextual fit, fidelity of implementation, and patient engagement.^60,61^ Downstream outcomes including pain, function, and quality of life may be delayed, diluted, or confounded by ineffectively applied interventions, local service constraints, and local changes in care as a result. In this sense, the impact of a guide may be better understood first through its influence on implementation processes and care delivery than through immediate changes in clinical endpoints alone.^62^ The BPG may be expected to have earliest influence on the consistency, acceptability, and context-sensitive delivery of care, with dissemination through educational offerings and an ‘adapt-not-adopt’ approach intended to maximise its relevance, uptake, and potential for improved patient outcomes over time.^63^

Related to the BPG formulation and educational packages, a participatory research approach may be warranted to ensure uptake in particular contexts. This work is in preparation and the protocol will be published separately.

## CONCLUSIONS

Best practice guides that meet the AGREE II criteria are needed to guide the management of people with a range of common musculoskeletal conditions to optimise care and improve outcomes. We have proposed, and initiated, a funded package of work to produce these guidelines with a comprehensive dissemination package to facilitate implementation.

## Data Availability

All data produced in the present study will be available upon reasonable request to the authors

## ACKNOWLEDGEMENTS

We would like to particularly acknowledge the patients, public, professionals and other stakeholders who contribute to the PPIEP activities and participate in this study. We would also like to acknowledge the funders for their unwavering support, QMUL colleagues in the joint research management office, and Gill Morrey for project management.

## FUNDING

Funding was provided by different organizations through multiple pathways.

The funders did not play a role in developing the protocol.

This work has primarily been supported by the Private Physiotherapy Educational Foundation Silver Jubilee Award (QMUL reference 13569244).

This work was supported by multiple small grants from Queen Mary’s Centre for Public Engagement.

Dissemination work has been supported by Queen Mary’s Impact Fund.

This work acknowledges the support of the National Institute for Health Research Barts Biomedical Research Centre (NIHR203330) for researchers employed at QMUL and Barts Health NHS trust.

This work acknowledges the support of Podium Analytics and the Podium Institute, a charity with the mission of More Sport Less Injury. https://podiumanalytics.org

## Notes

### Competing Interest Statement

The authors have declared no competing interest.

### Author Declarations

Ethical approval for the qualitative studies will be gained from Queen Mary University of London Research Ethics Committee for each condition separately. Ethics will also be obtained from the NHS to facilitate recruitment of patients with the selected conditions.

