## Appendix for "Data to Practice (D2P). Protocol for the development, dissemination and initial implementation of best practice guides for common musculoskeletal conditions: a mixed-methods study"

Appendix 1. Example of treatment classification into three levels for Rotator Cuff Tendinopathy.

| **First Level** | **Second Level** | **Third Level** |
| --- | --- | --- |
| 1. Minimal intervention | 1.1 Wait and See |  |
|  | 1.2 Inert Sham |  |
|  | 1.3 Sham |  |
| 2 Electrophysical | 2.1 Ultrasound |  |
|  | 2.2 Electrical Shockwave Therapy (ESWT) | 2.2.1 Focused ESWT |
|  |  | 2.2.2 Radial ESWT |
|  | 2.3 Laser/photobiomodulation |  |
|  | 2.4 Electrotherapy | 2.4.1 TENS |
|  |  | 2.4.2 Percutaneous Peripheral Nerve Stimulation |
|  |  | 2.4.3 NMES |
|  |  | 2.4.4 IFC |
|  |  | 2.4.5 Percutaneous Electrolysis |
|  |  | 2.4.6 TDCS |
|  | 2.5 Other electrophysical | 2.5.1 Diathermy |
|  |  | 2.5.2 Electromagnetic |
|  |  | 2.5.3 Infrared |
|  |  | 2.5.4 Radiofrequency |
| 3. Manual Therapy | 3.1 Spinal |  |
|  | 3.2 Local | 3.2.1 Glenohumeral /Scapulohumeral girdle |
|  |  | 3.2.2 Soft Tissue |
|  |  | 3.2.3 Muscle Energy Technique |
| 4. Taping | 4.1 Rigid Taping |  |
|  | 4.2 Elastic Taping |  |
| 5. Surgery | 5.1 Open Surgery |  |
|  | 5.2 Minimally Invasive Surgery |  |
| 6. Medication | 6.1 Oral medication |  |
|  | 6.2 Topical medication |  |
|  | 6.3 Supplements |  |
|  | 6.4 Medication unclear |  |
| 7. Needling | 7.1 Acupuncture |  |
|  | 7.2 Dry needling |  |
| 8. Education | 8.1 Information provision |  |
|  | 8.2 Formal Education |  |
| 9. Injection | 9.1 Injection – Steroid | 9.1.1 Local |
|  |  | 9.1.2 Systemic |
|  | 9.2 Non-Steroid Injection | 9.2.1 PRP |
|  |  | 9.2.2 Hyaluronic acid |
|  |  | 9.2.3 Botulinum toxin |
|  |  | 9.2.4 Glucose |
|  |  | 9.2.5 Anesthetic |
|  |  | 9.2.6 Ozone |
|  |  | 9.2.7 Tenoxicam |
|  |  | 9.2.8 Placenta |
|  |  | 9.2.9 Saline |
| 10. Exercise | 10.1 Strengthening | 10.1.1 High Intensity Well-Reported |
|  |  | 10.1.2 Low Intensity Well-Reported |
|  |  | 10.1.3 High Intensity Poorly-Reported |
|  |  | 10.1.4 Low Intensity Poorly-Reported |
|  |  | 10.1.5 Unclear Intensity Poorly-Reported |
|  |  | 10.1.6 Unclear Intensity Well-Reported |
|  | 10.2 Range of Motion (ROM) |  |
|  | 10.3 Movement Pattern Retraining (MPR) | 10.3.1 Motor Control |
|  |  | 10.3.2 Proprioception |
|  |  | 10.3.3 Stability |
| 11. Thermal | 11.1 Cryotherapy |  |
|  | 11.2 Heat |  |
| 12. Complex Intervention | 12.1 Complex Intervention | 12.1.1 Complex Intervention - Well Developed |
|  |  | 12.1.2 Complex Intervention - Poorly Developed |
|  | 12.2 Impairment-based Complex Intervention |  |
| 13. Other Multimodal Treatment | 13.1 Highly-Complicated Multi-treatment Interventions |  |
|  | 13.2 Unclear Complex Intervention |  |
| 14. Other | 14.1 Ultrasonophoresis |  |
|  | 14.2 Graded Motor Imagery |  |
